# Using Retinal diagnostics as a Biomarker for Neurodegenerative Diseases: Protocol for a systematic review

**DOI:** 10.1101/2024.05.27.24306765

**Authors:** Zeynep Sahin, Sara Pisani, Paul Nderitu, Ashwin V Venkataraman, Ta-Wei Guu, Dag Aarsland, Timothy Jackson, Dominic ffytche

## Abstract

**Introduction:** Retinal neurodegeneration has recently been shown to occur in tandem with neurodegenerative disease. In the expectation that disease modifying treatments for Alzheimer’s Disease and Parkinson’s Disease will soon become available, it will be important to have clinically useful biomarkers for neurodegenerative disease subtyping to guide early diagnosis, inform on prognosis and stratify subgroups for treatment. Understanding differences in detectable retina changes in individuals with different neurodegenerative disease subtypes is therefore fundamental. The emerging field of oculomics posits that systemic and neurodegenerative disease can be characterised using detectable ocular biomarkers within retinal diagnostics. The aim of this review is to compare the performance of common retinal imaging modalities in neurodegenerative disease detection and subtyping.

**Methods and analysis:** This protocol has been developed in accordance with the *Preferred Reporting Items for Systematic Reviews and Meta-Analyses Protocols* (PRISMA-P) guidelines. A comprehensive literature search will be conducted in PubMed, Web of Science, and Scopus. Eligible studies will have reported using retinal diagnostic tools defined as Optical Coherence Tomography (OCT), Optical Coherence Tomography Angiography (OCTA), Colour Fundus Photography (CFP) and Electroretinography (ERG) in individuals with neurodegenerative diseases, including Alzheimer’s Disease (AD), Parkinson’s Disease (PD), Dementia with Lewy Bodies (DLB), Frontotemporal Dementia (FTD), Vascular Dementia (VaD), and Mild Cognitive Impairment (MCI). There will be no time restrictions placed in these searches. Studies not written in English, not peer-reviewed and grey literature will be excluded. Screening for eligible studies and data extraction will be conducted by two independent reviewers, using predefined inclusion criteria. Any disagreements between the reviewers will be settled by discussion, and if required, third senior reviewer arbitration. The systematic review primary outcome is the performance of retinal diagnostics, namely OCT, OCTA, CFP, and ERG in the detection and subtyping of aforementioned neurodegenerative diseases. The secondary outcome is to evaluate the association between changes in retinal diagnostic features (e.g. retinal layer thicknesses) and neurodegenerative disease subtypes. The quality of the included studies will be assessed using the Grading of Recommendations, Assessment, Development and Evaluations (GRADE) tool. A narrative synthesis approach will be used to analyse the results, with meta-analysis performed if there is sufficient data.

**Ethics and Dissemination:** Ethical approval for this manuscript is not required, as this is a protocol for a systematic review and therefore no data are to be collected. Findings for this systematic review will be disseminated as a peer-reviewed publication and presentations at national and international symposiums including International Lewy body Dementia Conference, International Congress of Parkinson’s Disease and Movement Disorders, The Association for Research in Vision and Ophthalmology.

**PROSPERO Registration Number:** CRD42023434024

**STRENGTHS AND LIMITATIONS OF THIS STUDY:**

▪ Our aim is to perform a comprehensive systematic review of the performance of retinal diagnostic methods, namely OCT, OCTA, CFP, and ERG in neurodegenerative disease subtyping.
▪ We will use the carefully defined methodology in accordance with the Cochrane handbook, and the results of this systematic review will be reported as per Preferred Reporting Items for Systematic Reviews and Meta-Analyses Protocol (PRISMA-P) statement.
▪ The certainty of this systematic review may be limited due to the small sample of studies for Dementia with Lewy Bodies (DLB), Frontotemporal Dementia (FTD), and Vascular Dementia (VaD).

## BACKGROUND

*Neurodegenerative disease* is an umbrella term used to describe a wide array of neurological conditions resulting in loss of nerve cells and gradual decline brain function. Alzheimer’s Disease (AD) and Parkinson’s Disease (PD) are amongst the most diagnosed neurodegenerative diseases in the United Kingdom, affecting 1 million people^1^. According to the Office for National Statistics, Dementia and AD was the leading cause of death, per 100,000 people, in England in December 2020 on a 5-year average^2^, and is continuing to increase in prevalence in the UK^3^. There is an urgent need not only for disease modifying treatments^4^, but also timely and specific diagnostics for this cluster of diseases.

While not commonly conceptualised in such terms, the eye is an anatomical extension of the brain and central nervous system. The retina functions as a sensory structure responsible for transducing luminous stimuli into electrical signals. Subsequently, these electrical signals traverse a ganglion nerve cells to the optic nerve and ultimately reaching the visual cortex through a series of synaptic connections within the optic nerve pathway. Due to the similarities between the retina and brain in terms of cell composition, structure, and the inflammatory and immunologic reactions to insults^5,6^ embryological extension of the brain with shared cell types^7^, the eye offers a unique window into brain neurodegenerative changes as retinal changes can reflect the same pathology as the brain^8^. Recent developments in retinal diagnostics have highlighted the eye as an important indicator of systemic health, an area of study newly coined as *oculomics*^9^. Subtle changes detectable on retinal imaging are associated with risk of many leading causes of morbidity and mortality, such as heart diseases^10^, stroke^11^, and neurodegenerative diseases^12^. Hence, evidence from studies of retinal imaging can provide valuable insights into the early detection of neurodegenerative diseases^13^. Structural retinal measures derived from optical coherence tomography (OCT) imaging have been shown to differ in established AD and PD, with some studies also looking into Vascular Dementia (VaD), Frontotemporal Dementia (FTD) and Dementia with Lewy Bodies (DLB).

Increasing evidence is suggesting that retinal imaging changes could be an imaging biomarker for neurodegenerative diseases, and that thinner retinal nerve fibre (RNFL), ganglion cell layers (GCL), inner plexiform layers (IPL) and inner nuclear layer (INL)^12^ can reflect global brain atrophy and the development of dementia. However, little is known about the association between changes in retinal cell layers with respect to the differing sub-types of dementia to guide which imaging modality is best suited to detect these changes. As well as identifying the gold-standard retinal imaging approach which can be used as a new biomarker for the detection of neurodegenerative diseases, our aim is also to identify which of the intricate cellular layers are affected with different diseases. Retinal diagnostics are accessible, non-invasive, and cheaper methods compared to neuroimaging modalities used for diagnosis presently. Therefore, retinal diagnostic methods could have a potential utility in the detection of neurodegenerative diseases and importantly for disease subtyping to guide earlier diagnosis and treatment. Thus, this systematic review could have positive health and diagnostic outcomes if there are differences in the involvement of retinal cell layers in sub-types of neurocognitive diseases detectable with retinal diagnostic tools.

## METHODS AND ANALYSIS

### Study Design

A comprehensive literature search of electronic bibliographic databases will be conducted on PubMed, Web of Science and Scopus. There will be no limits on publication year applied. However, only studies which have been peer-reviewed, published in English and are original research publications will be included in the systematic review. Furthermore, reference lists of identified studies, systematic reviews and meta-analyses will be scoped for additional sources to search for further studies that may have not appeared in the original database search. Results from these databases will be merged using an electronic reference manager to facilitate removal of duplicates. This systematic review will follow the guidelines recommended by the updated Preferred Reporting Items for Systematic Reviews and Meta-Analyses (PRISMA-P).

### Participants

Inclusion criteria for this systematic review will be mid-to-late-life adults aged ≥50 with AD, DLB, PD, FTD, VaD or MCI as defined by the World Health Organisation (WHO) International Classification of Diseases 11^th^ Revision (ICD-11), as well as mid-to-late-life healthy adults aged ≥50 with Subjective Cognitive Disorder (SCD) and cognitively healthy controls with and without family history of neurodegeneration.

### Eligibility

Different subtypes of neurocognitive diseases will be compared with cognitively healthy controls or other neurodegenerative conditions. Comparison of the different subtypes will also be made to investigate the possible dynamic retinal changes. Furthermore, studies describing at least one type of retinal diagnostic tool will be included in this systematic review. Retinal diagnostic modalities include, but are not limited to, colour fundus photography (CFP), Optical Coherence Tomography (OCT), Optical Coherence Tomography Angiography (OCTA), and Electroretinography (ERG). Population-based cohorts will be considered if a neurocognitive assessment, such as Mini-Mental State Examination (MMSE), Montreal Cognitive Assessment (MoCA), Addenbrooke’s Cognitive Examination (ACE), has been administered. Reviews, case studies, and animal studies will be excluded from this systematic review.

### Outcomes

The primary outcome of this systematic review would be to understand (1) whether different high-resolution retinal imaging techniques can detect neurodegenerative disease based on changes in the retina.

Secondary outcome is (2) to investigate which retinal layers or structures are affected in different sub-types of neurodegenerative disease. This will be defined by the presence of the following features where available:

CFP:

▪ Retinal vascular morphology
  ○ Central retinal artery and vein equivalents
  ○ Fractal dimension
  ○ Vessel density
  ○ Vesel widths
▪ Optic nerve head morphology
  ○ Cup to disc ratios

OCT (Macula):

▪ Retinal thickness measures
  ○ Macula RNFL
  ○ GCL
  ○ Inner Plexiform Layer (IPL)
  ○ GCIPL (GCL + IPL)
  ○ Inner Nuclear Layer (INL)
  ○ Outer Plexiform Layer (OPL)
  ○ Outer Nuclear Layer (ONL)
  ○ Total retinal thickness

OCT (Peripapillary RNFL):

▪ pRNFL
  ○ Sector thickness
  ○ Global thickness

OCTA:

▪ Foveal Avascular Zone (FAZ)
▪ Superficial Vascular Plexus (SVP) density / fractal dimension
▪ Deep Capillary Plexus (DCP) density / fractal dimension

ERG:

▪ Full field amplitude / implicit time
▪ Flicker peak to peak time / amplitude
▪ Multifocal electroretinogram (mfERG) amplitudes, implicit times, and ring ratios

Furthermore, if available, data on Apolipoprotein-E-(ApoE) genotyping, Amyloid-β (Aβ), Alpha-synuclein (α-Syn) from cerebrospinal fluid (CSF) and neuroimaging modalities used will also be extracted.

### Study selection

A search for Web of Science has been provided as an example and is available in online supplemental appendix 1. The search terms used were:

▪ “Alzheimer’s Disease”, “Neurodegeneration”, “Mild Cognitive Impairment”, “Frontotemporal Dementia”, “Subjective Cognitive Impairment”, “Vascular Dementia”
▪ “Retinal Imaging Dementia”, “Retinal Imaging”, “Optical Imaging”, “Ocular Imaging”

One researcher will independently conduct a literature scope. Two researchers will then independently screen titles and abstracts for the study selection process and extract the relevant data. Any disagreements will be settled by consensus and discussion. If required, a senior moderator will arbitrate.

### Study extraction

Data extracted by the two researchers from full text articles include study design, demographics, clinical diagnosis, sample size, sample groups, neurocognitive tests used, and disease subtype. For each of the retinal imaging modalities, we will extract data on retinal vascular morphology, retinal cell layer thicknesses and/ or retinal functional measurements determined from the ERG. After data extraction, the two researchers will confer and any disagreements will be settled by consensus and discussion with senior moderator will arbitration when necessary.

### Risk of bias assessment

The quality assessment will be conducted by the same two independent researchers who will complete the extraction process, and they will be using the critical appraisal tool for specifically assessing risk of bias in prevalence studies. Overall, this tool includes nine-questions scoring 0-1 (0 low risk, 1 high-risk) to determine bias in data analysis and selection bias. Any disagreements will be resolved by discussion or referral to a third (senior) reviewer.

### Data analysis

The data collected for this review will be analysed using a purpose-built software for systematic reviews and meta-analysis called ReviewManager (RevMan). Studies which meet the eligibility criteria will be divided into categories according to disease subtype. Characteristics of included studies will be presented in summary tables and narrative text. Studies focusing on more than one subtype of neurodegenerative disease will be treated as independent studies. For example, a study including AD, PD and healthy controls will be categorised under the AD domain and PD domain. Neurodegenerative disease will be divided into subgroups to assess whether different retinal diagnostic techniques can detect the different subtypes of neurodegenerative diseases, as well as which of these retinal imaging modalities would be the most effective in disease detection. As this manuscript will be published as a systematic review, a narrative synthesis will be chosen to display the results.

### Grading of evidence

Quality of the data extracted will be assessed using the Grading of Recommendations, Assessment, Development and Evaluations (GRADE) tool^14^. Manuscripts will be considered to constitute high-certainty evidence to answer the systematic review question and will be downgraded for risk of bias, inconsistency, indirectness, imprecision, and publication bias. Two reviewers will independently make this decision. If necessary, a third senior review will arbitrate.

### Patient and public involvement

There were no time and funds allocated for a patient and public involvement event. However, this systematic review will be asking an important clinical question, and the protocol follows a standardised approach as per PRISMA guidelines. A Patient and Public Involvement and Engagement (PPIE) group will be convened to disseminate findings once the review has been completed.

## ETHICS AND DISSEMINATION

As this is a systematic review using published data without the need of accessing personal identifiable information, there are no immediate ethical or safety concerns. The results from this systematic review will be submitted for publication in a peer-reviewed open-access journal, disseminated to relevant dementia charities, and will be presented at national and international meetings.

## CONCLUSION

With the increase of burden of the diseases on the individual and families there is an urgent need for quicker, cheaper, and robust imaging biomarker for a timely diagnosis. This systematic review will gather and disseminate information to see whether using commonly available retinal diagnostic methods will be able to detect, and potentially subtype, neurodegenerative diseases accurately in a clinical use.

## Data Availability

All data produced in the present work are contained in the manuscript

