## appendix 1 for "Using Retinal diagnostics as a Biomarker for Neurodegenerative Diseases: Protocol for a systematic review"

Appendix 1. SEARCH STRATERGY FOR WEB OF SCIENCE

**#9 AND #14** and **English** (Languages) and **Article** (Document Types) and **Article** (Document Types) and **Human** (Search within all fields) and **Article** (Document Types)

**#9 AND #14** and **English** (Languages) and **Article** (Document Types) and **Article** (Document Types) and **Human** (Search within all fields)

**14. #10 OR #11 OR #12 OR #13**

**13. ocular imaging** (All Fields)

**12. optical imaging** (All Fields)

**11. retinal imaging** (All Fields)

**10. retinal imaging dementia** (All Fields)

**9. #1 OR #2 OR #3 OR #4 OR #5 OR #6 OR #7 OR #8**

**8. vascular dem*** (All Fields)

**7. subjective cognitive impair*** (All Fields)

**6. subjective cognitive dis*** (All Fields)

**5. frontotemporal dem*** (All Fields)

**4. mild cognitive impair*** (All Fields)

**3. neurodegen*** (All Fields)

**2. alzheimer*** (All Fields)

**1. dementia** (All Fields)
